# Green Heart Louisville: intra-urban, hyperlocal land-use regression modeling of nitrogen oxides and ozone

**DOI:** 10.1101/2023.03.03.23286765

**Authors:** Pradeep Prathibha, Raymond Yeager, Aruni Bhatnagar, Jay Turner

## Abstract

Exposure to urban air pollution is linked to increased mortality from cardiopulmonary causes. Urban areas juxtapose large numbers of residences and workplaces with near-road environments, exacerbating traffic-related air pollution (TRAP) exposure. TRAP is the primary source of variability in intraurban air quality, but continuous regulatory monitoring stations lack the spatial resolution to detect fine-scale pollutant patterns that recent studies using long-term, resource-intensive mobile measurements have established as persistent and associated with higher risk of cardiovascular events. This work evaluates a low-cost, fixed-site approach to characterizinglong-term, hyperlocal exposure to oxides of nitrogen (including NO_2_, a common surrogate for TRAP) as part of Green Heart Louisville, a prospective cohort study examining linkages between urban vegetation, local air quality, and cardiovascular health.

We used a fixed 60-site network of Ogawa passive samplers in a 12 km^2^ section of Louisville, KY, to measure two-week integrated NO_2_, NO_x_ (NO + NO_2_), and O_3_ mixing ratios nominally every two months between May 2018-March 2021. Seasonal NO_x_ averages were 2.5-fold higher during winter than in summer, and annual average NO (calculated by difference in NO_x_ and NO_2_) and NO_2_ ranged from 4-21 ppb and 5-12 ppb, respectively. NO increased 3-to-5-fold within 150 m of highways or major arterial roads and 2-to-3-fold near parking lots. While both NO and NO_2_ were elevated in near-road environments, the corresponding O_3_ was depressed, consistent with titration by NO.

We developed land-use regression models for annual average NO, NO_2_, and NO_x_ using parameters of proximity (distance to nearest road type, restaurant, traffic signal), cumulative occurrence (length of roads, number of restaurants and traffic lights, all in buffers of up to 500 m in 50-m increments), and greenness (normalized difference vegetative index (NDVI)). Adjusted spatial variability explained by the models were 70% (p<0.05), 67% (p<0.05), and 75% (p<0.01) for NO, NO_2_, and NO_x_, respectively. Common predictors were distances to the nearest restaurant and road as well as total length of roads within 350 m. Only one greenness metric was significant: mean NDVI within 50 m was negatively associated (p=0.02) with NO_2_. We plan to use these hyperlocal models to estimate residential-level exposures of the clinical study participants.

## Introduction

Epidemiological and toxicological studies have linked urban air pollution exposure to adverse health outcomes ranging from minor respiratory irritation to chronic cardiometabolic diseases and an increase in natural-cause mortality (1–3). Urban areas, projected to expand by up to 1.3 million km^2^ in 2050 to accommodate 70% of a global population of 10 billion, juxtapose large numbers of residences and workplaces to near-road environments (4,5). Traffic-related air pollution (TRAP) is elevated beyond urban background levels in these environments and is the primary source of variability in intraurban air quality; several studies indicate this variation exceeds interurban variations (6–11).

Numerous studies have explored the associations between health effects and exposure to such spatially heterogeneous air quality using pollutant concentrations measured at central monitoring sites. The ambient air quality monitoring network in the United States consists of ∼4000 sties, each with a zone of representation of 1 km^2^, averaging to ∼2-5 monitoring stations per million people and 1000 km^2^ (12–14). The low spatial resolution of such data can misclassify exposures and obscure finer-scale associations to health effects (15). This deficit has been addressed in recent years through small-area, fixed-site network measurements due to advances in both geospatial analytical tools and low-cost measurement technologies (16,17). In particular, landuse regression (LUR) modeling has been employed extensively to predict intraurban concentrations of TRAP components (15,18–22). LUR models treat pollutant concentration as the dependent variable, with a linear combination of the characteristics of the surrounding natural and built environment as the independent variables. These characteristics are quantified by Euclidean proximity or in buffers of varying radii around the site of measurement, and typically include traffic volume and lengths of various road types, population or residential density, land use types (residential, commercial, open areas, etc.), and wind direction in relation to major point sources (23,24). As demonstrated by Briggs *et* al. in the first study to use LUR modeling to predict air quality, this approach is empirical and thus, area-specific: applying a model developed for one city to another failed to evince pollutant spatial heterogeneity (23).

Numerous area-specific studies in North America and Europe report robust models to estimate exposure to intraurban pollution using nitrogen dioxide (NO_2_), a good indicator for TRAP (25,26). LUR models for NO_2_ based on fixed-site measurements produce good predictions (0.46 < R^2^ < 0.94), but are spatiotemporally limited (8,27–35). Networks reported to date have nominally used 1 site per km^2^, with a highest reported density of 3 sites/km^2^ in Amsterdam for the SAVIAH Project (23). Temporal coverage is also limited, with each study conducting 1-4 deployments. This spatial resolution falls short of the scales sought by assessments of finer-scale health effects: intraurban TRAP has been noted to vary over the scale of 50 m or less (36,37).

Given this gap in hyperlocal, temporal assessments necessary to assess associations with fine scale health effects, we measured NO_2_, nitric oxide (NO, a primary pollution in vehicular exhaust that rapidly oxides to NO_2_), and ozone (O_3_) mixing ratios (henceforth referred to as concentrations) nominally every two months between May 2018-March 2021 in a dense (5 sites/km^2^) network in Louisville, KY. Then, we developed LUR models fitted to land use and land cover to determine whether the fixed-site approach can capture long-term, hyperlocal patterns in NO_x_ and O_3_.

## Methods

### Study area

The passive sampling network spanned a 12 km^2^ portion located ∼5 km east of the Ohio River and with a population of 25,000 in metropolitan Louisville (38° 11’ 38”, -85° 46’ 29”). The study area (Figure 1), chosen for a prospective cohort study of cardiovascular disease risk and neighborhood level greenness (Green Heart Louisville), is bisected from east to west by a major interstate highway (I-265, Watterson Expressway), is flanked along the northwest and east by freight railway lines, and is adjacent to an international airport that serves as the worldwide cargo hub of a major air freight company. While land use within the study domain is mostly residential with some commercial spaces along major roads, that outside the perimeter is commercial, industrial, or public space used for parking and other transportation use.

**Figure 1.**
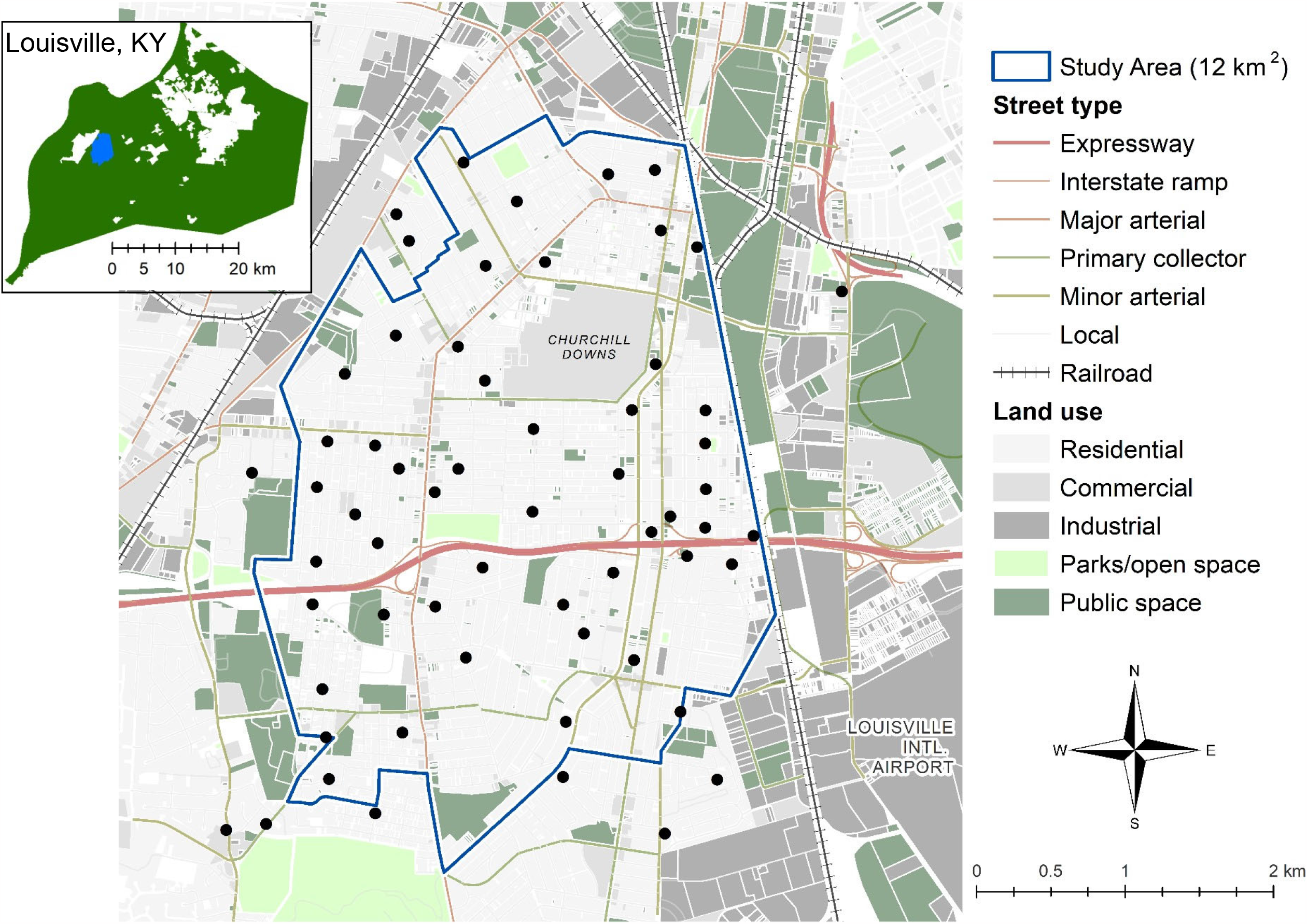
Land use within the study domain and the 60 fixed sites (black circles) in the passive sampling network in metropolitan Louisville, KY (inset, with the study domain shaded blue).

### Sampling network and method

The location-allocation method was used to identify 60 spatially balanced points weighted by population density and NO_2_ concentration from a satellite-derived exposure surface (Su 2018). For each point, the nearest accessible utility pole was chosen for passive sampling (Figure 1), and the location of each pole was mapped using BT-Q1000XT global positioning system (Qstarz International Co. Ltd., Taiwan, R.O.C.). Badge passive samplers for NO_x_ and O_3_ (Ogawa & Co., USA) were assembled in St. Louis, shipped to Louisville in coolers with ice packs, and stored in - 4°C freezers until deployment. A field technician deployed the samplers at a height of 2 m for 14-day sampling. Each site had two dual-sided samplers, one with NO_2_ and NO_x_ sampling pads, and the other with one O_3_ sampling pad. The samplers were deployed and retrieved over two days, with the time of installation and retrieval noted at every site. We used ion chromatography to quantify adsorbed NO_2_, NO_x_, and O_3_ species and determined ambient concentrations using protocols by Ogawa (NO-NO_x_, NO_2_, SO_2_ sampling – v6.0, O_3_ measurement using the O_3_ passive sampler badge – v3.0); NO concentrations were calculated by the difference in NO_x_ and NO_2_. Concentration conversion coefficients were calculated using hourly weather data monitored at the Louisville International Airport station (KSDF) retrieved from Weather Underground (www.wunderground.com).

### Quality control

For each pollutant, collocated precision was determined using collocated samples collected at the same six, spatially distributed sites in every deployment. Three field blanks were used to quantify contamination due to transport and ambient conditions and one equipment blank to track laboratory contamination. Samplers that appeared to have been disturbed (shelters missing or samplers unclipped from brackets) were excluded from analysis. We compared NO_2_ and O_3_ concentrations averaged by deployment with contemporaneous data from the nearest regulatory monitoring site (∼12 km away, Cannon’s Lane (38° 13’ 43”, -85° 39’ 16”)) using Pearson’s correlation coefficient (ρ). We also collected collocated samples (N=3) at the regulatory monitoring site for direct comparison.

### Predictor variables

We obtained street centerline (2020), railway (2019), land use (2019), food service locations (2021), and signalized intersections (2021) datasets from the City of Louisville’s open data portal. We also identified airports, manufacturing complexes, and educational complexes using Areas of Interest (2021) (39). Streets were reclassified into expressways, interstate ramps, primary collectors, major arterials, minor arterials, and local roads, and land use polygons were classified into residential, commercial, industrial, public spaces, and parks or open spaces.

To quantify greenness, we used NDVI (normalized difference vegetation index) computed from 8-bit 3-band (RGB) orthoimagery acquired in a single aerial lift on September 9, 2019, as well as leaf biomass and leaf area. Tree-specific values for the two latter parameters were predicted using a deep learning convolutional predictive model to identify individual trees from aerial imagery. R^2^ between predicted and actual (ground-truth) values were 0.65 and 0.57 for leaf biomass and leaf area, respectively, and buffer-specific means were normalized to a unitless range (0-1) by dividing by the maximum value. The collection and processing of greenness datasets were completed by collaborators.

Then, at each sampling location, we used ArcGIS 10.6.1 (ESRI®, Redlands, CA) to compute variables by 1) proximity (the nearest road by type, the nearest potential emission source (food service locations that prepare and not just vend food, school cafeterias, parcels classified for manufacturing use), or feature that could aggregate vehicles (traffic signals, parking lots)) and 2) cumulative occurrence (total length of roads by type, number of restaurants and traffic signals, total area of each land use type, fraction of area of each land use type, average NDVI/leaf biomass/leaf area) in buffers up to 500 m in 50 m increments. We crudely classified 7-inch NDVI raster into three classes to distinguish paved or roof surfaces (−1–0.17), grasses (0.17–0.42), and larger vegetation (brushes, forbs, and trees; 0.42–0.8); we only used the latter class to compute buffer averages. In all, this generated 159 independent variables.

### Model development and validation

All regression analyses and diagnostics were conducted in R version 3.6.3 (The R Foundation for Statistical Computing, www.r-project.org). As the focus is to detect spatial heterogeneity in pollutant concentrations, for each pollutant, we background-adjusted the data by subtracting the 10^th^ percentile of annual average across all sites; this value was added to model predictions for visualizations. Note that this will only affect the fitted intercept and not the significances or magnitudes of coefficients in the model. Due to the high collinearly expected in the independent variables, backward stepwise selection was used to determine the combination of predictors that explained the most variance in pollutant concentrations, as quantified by the adjusted coefficient of determination (adjusted R^2^). The most collinear variable, identified through the highest variance inflation factor (VIF), was eliminated per iteration if the adjusted R^2^ remained unchanged (±5%) upon its exclusion. Once the VIF were within 3-5 points, parameters with the highest p-values (only if p>0.05) were eliminated until all retained predictors had VIF≤5. We chose this threshold, which is more liberal than those employed in some larger-scale LUR studies (VIF<2), to accommodate the higher homogeneity inherent to the smaller spatial extent. For each pollutant, I first constructed separate models with just parameters of proximity and parameters of cumulative occurrence; then, we combined the significant terms into a single model and repeated the elimination process. Final models were validated using leave-one-out-cross-validation (LOOCV), in which models are applied to n–1 sites (in this case, 59) to compare predicted and measured concentration at the left-out site; this was repeated for each of the sites in the network. We chose this validation method because it uses each observation for both training and validation, suppressing variability that will otherwise be high for datasets of this size. The final models were also applied to seasonal average concentrations (summer and winter only, background-adjusted by 10^th^ percentile of the season-specific average) to assess whether the predictions remained significant at smaller temporal scales.

## Results

### Independent variables: Ambient NO_x_ and O_3_

19 full-scale deployments have been conducted to date, but for all analyses in this study, we used data from 16 NO_x_ and 14 O_3_ deployments in 2018-2020 (Table 1) as contemporaneous data were available from the regulatory monitoring site for validation. Samplers from at most three sites were missing or otherwise compromised during each run, and within the same year, no location had more than one value missing; because of this, missing values were not imputed. Minimum detection limits (MDL) of ambient mixing ratios were 0.45 ppb for NO_2_, 3.81 ppb for NO_x_, 3.36 ppb for NO, and 4.2 ppb for O_3_ (40 No concentrations from field-deployed samplers were below these MDLs. However, effective ambient concentrations from field and laboratory blanks were below MDL for nearly two-thirds of the measurements, and we imputed these values with the corresponding MDL.

**Table 1.** Summary of 2-week sampling of NO_x_ and O_3_ in the 60-site network

| Deployment | Start date <sup>a</sup> | End date <sup>a</sup> | Season |
| --- | --- | --- | --- |
| 1<sup>b</sup> | May 15-16, 2018 | May 29-30, 2018 | Summer |
| 2<sup>b</sup> | Jul 11-12, 2018 | Jul 25-26, 2018 | Summer |
| 3 | Sep 26-27, 2018 | Oct 10-11, 2018 | Fall |
| 4 | Jan 8-9, 2019 | Jan 22-24, 2019 | Winter |
| 5 | Mar 6-7, 2019 | Mar 20-21, 2019 | Spring |
| 6 | May 21-22, 2019 | Jun 4-5, 2019 | Summer |
| 7 | Jul 11-12, 2019 | Jul 25-26, 2019 | Summer |
| 8 | Sep 9-10, 2019 | Sep 23-24, 2019 | Fall |
| 9 | Nov 20-21, 2019 | Dec 3-4, 2019 | Winter |
| 10 | Mar 16-17, 2020 | Mar 30-31, 2020 | Spring |
| 11 | Apr 30-May 1, 2020 | May 12-13, 2020 | Summer |
| 12 | Jun 3-4, 2020 | Jun 17-18, 2020 | Summer |
| 13 | Jul 8-9, 2020 | Jul 22-23, 2020 | Summer |
| 14 | Aug 26-27, 2020 | Sep 9-10, 2020 | Fall |
| 15 | Oct 6-7, 2020 | Oct 20-21, 2020 | Fall |
| 16 | Nov 18-19, 2020 | Dec 2-3, 2020 | Winter |

<sup>a</sup> Both deployment and retrieval completed over two days
<sup>b</sup> Only NO<sub>x</sub> measurements conducted during these runs; also, these samples were analyzed using colorimetric methods adapted from Ogawa's protocol (NO-NO<sub>x</sub>, NO<sub>2</sub>, SO<sub>2</sub> sampling – v6.0)

Annual average NO and NO_2_ ranged from 4-21 ppb and 5-12 ppb, respectively. Both were elevated in near-road environments, with NO increasing 3-to-5 fold within 150 m of highways and major arterial roads and 2-to-3 fold near parking lots; the corresponding O_3_ at these locations was depressed, which is characteristic of NO titration. Values at no location exceeded the National Ambient Air Quality Standards annual mean NO_2_ (53 ppb) or 3-year average fourth-highest daily maximum 8-hour O_3_ (70 ppb), but note that these 2-week samples lack the temporal resolution to be directly compared to O_3_ standard based on the annual fourth-highest daily maximum 8-hour concentration averaged over 3 years. Collocated precision calculated from six pairs in each deployment were 1.4 ppb for NO_2_, 2.1 ppb for NO_x_, 2.2 ppb for NO, and 2.9 ppb for O_3_.

Finally, collocated passive sampling of three NO_2_ and O_3_ at the regulatory monitoring station showed that two-week averages measured by the samplers were lower by 3.5 ppb and 1.8 ppb, respectively. However, comparing deployment-wide mean concentrations (Figure 2) showed that passive sampled mixing ratios (NO_2_: 8.1 ppb, O_3_: 29.2 ppb) were similar to the monitoring data (NO_2_: 8.2 ppb, O_3_: 28.9 ppb) and highly correlated for both NO_2_ (ρ=0.86, 95% CI 0.61, 0.95) and O_3_ (ρ=0.94, 95% CI 0.83, 0.98).

**Figure 2.**
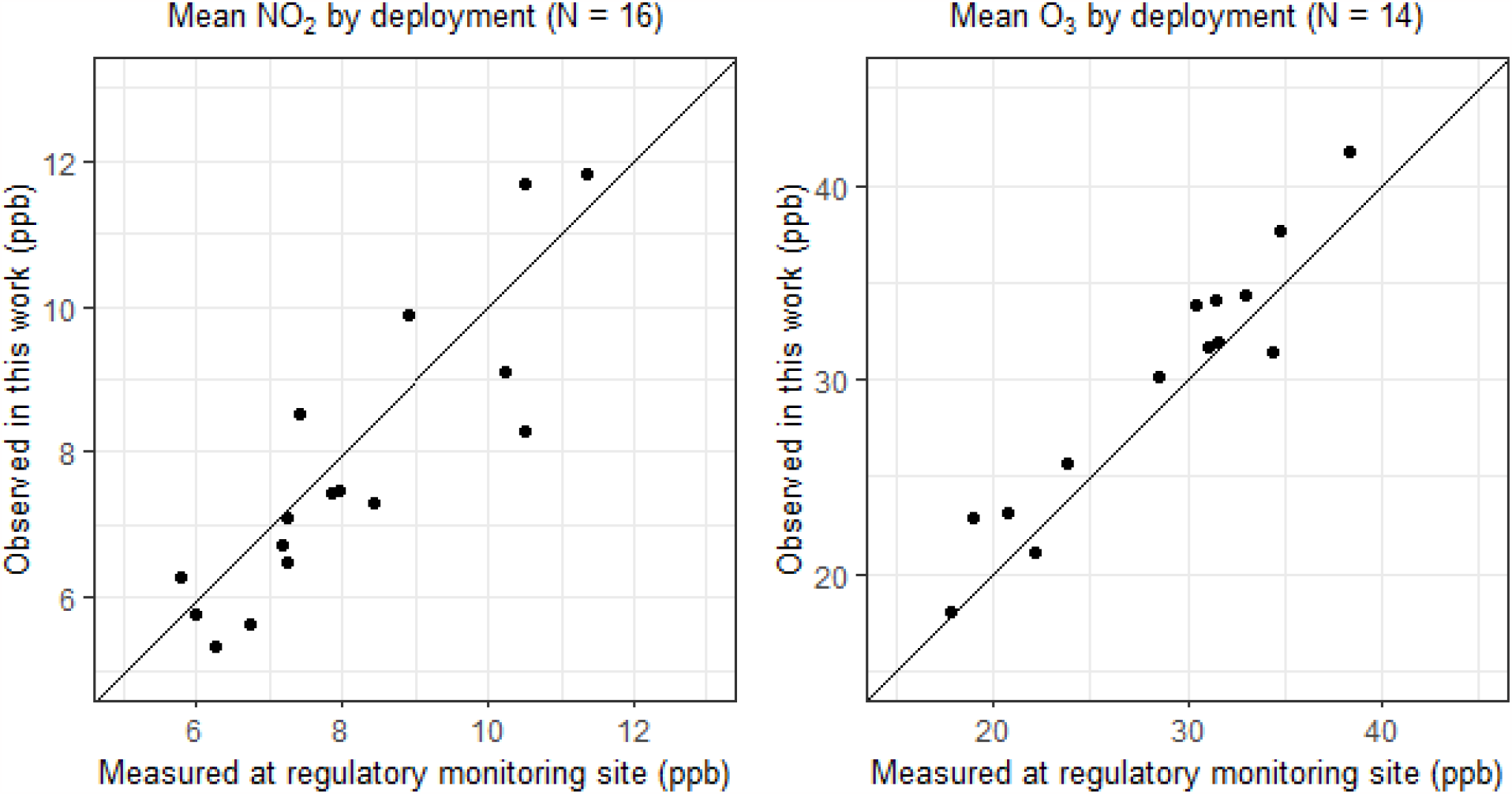
Comparison of 2-week mean concentrations measured in this study and at the nearest (∼12 km) regulatory monitoring site (Cannon’s Lane)

### Dependent variables: land-use characteristics

The most dominant land use feature was residential space, with single-family parcels occupying nearly 50% of the area across all buffer sizes; this changed to either public space or industrial and commercial areas for sites along the periphery of the study domain. The buffer average of NDVI ranged from 0-0.37 in all buffer sizes, and 25% of the sites were not within 500 m of any restaurants or food service locations. Leaf area per tree ranged from 12 m^2^ to 3,490 m^2^, while leaf biomass per tree ranged from 1 kg to 419 kg, corresponding to normalized buffer averages of 0.12 and 0.38, respectively. The street type most abundant in length and closest to all except one location was local road, and throughout the study domain, predictors characterizing traffic were highly correlated.

### Modeling results

The final regression models (Figure 3; fully tabulated in Table 2, Supplementary material) explained the spatial variation in annual average concentrations of NO by 66%, NO_2_ by 61%, NO_x_ by 71%, and O_3_ by 53%. Regression diagnostics revealed that residuals were normally distributed, and while all four models had three outlying standard residual values, all were within the Cook’s distance, suggesting they were not influential points in the regression line. Moreover, these points did not belong to the same location, and their exclusion only improved the adjusted R^2^ of the corresponding models modestly (<5%). There were no differences in models developed using annual average concentrations from 2019 or 2020 (adjusted R^2^ within <3%); however, summer and winter average concentrations correlated poorly (0.2<ρ<0.4) with predicted concentrations. Finally, LOOCV determined average differences of 0.5-1.0 ppb for NO, NO_2_, and NO_x_ models and 2.0-3.5 ppb for O_3_, both corresponding to average differences of 8-15%.

**Figure 3.**
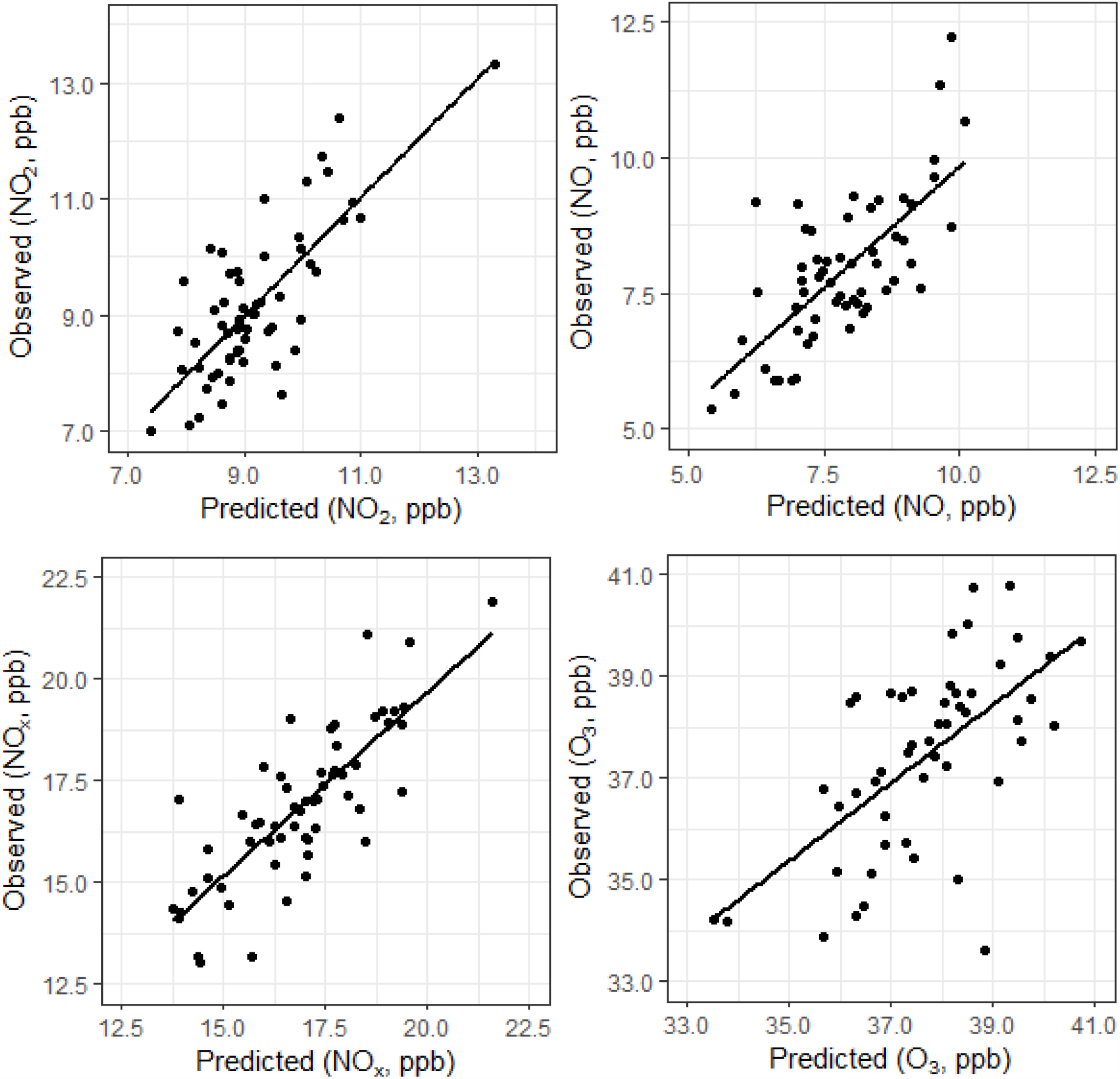
Comparison of predicted and measured annual average concentrations of NO, NO_2_, NO_x_, and O_3_.

**Table 2.** Summary of multivariable predictive models for annual average NO, NO_2_, NO_x_, and O_3_ (ppb) with standardized coefficients

| Type | Predictor | Coefficient | Standard error | t | Prob>t | VIF |
| --- | --- | --- | --- | --- | --- | --- |
| **NO (adj. R<sup>2</sup> = 0.66, p<0.001)** |  |  |  |  |  |  |
|  | Constant | 0.02 | 0.15 | 0.16 | 0.873 |  |
| Total length | Highway (<100 m) | 0.66 | 0.19 | 3.52 | 0.001 | 1.53 |
|  | Local (<250 m) | 0.44 | 0.20 | 2.22 | 0.032 | 1.76 |
|  | Minor arterial (<350 m) | 1.56 | 0.40 | 3.93 | 0.000 | 1.94 |
|  | Major arterial (<500 m) | 1.27 | 0.39 | 3.23 | 0.002 | 1.81 |
| Count | Restaurants (<250 m) | 0.57 | 0.27 | 2.10 | 0.042 | 3.24 |
|  | Restaurants (< 350 m) | 0.93 | 0.31 | 3.05 | 0.004 | 4.14 |
| Area | NDVI (50 m) | -1.29 | 0.35 | -3.71 | 0.001 | 3.35 |
| Proximity | Major arterial | 0.79 | 0.27 | 2.88 | 0.006 | 3.26 |
| **NO<sub>2</sub> (adj. R<sup>2</sup> = 0.61, p<0.001)** |  |  |  |  |  |  |
|  | Constant | -0.06 | 0.12 | -0.48 | 0.636 |  |
| Total length | Local (<500 m) | 0.38 | 0.15 | 2.64 | 0.011 | 1.52 |
|  | Major arterial (<100 m) | 0.46 | 0.13 | 3.48 | 0.001 | 1.24 |
|  | Minor arterial (<100 m) | 0.37 | 0.15 | 2.44 | 0.018 | 1.64 |
|  | Minor arterial (<500 m) | 0.29 | 0.15 | 1.93 | 0.059 | 1.56 |
|  | Primary collector (<500 m) | 0.38 | 0.13 | 2.88 | 0.006 | 1.23 |
|  | Ramp (<100 m) | 0.52 | 0.16 | 3.27 | 0.002 | 1.83 |
|  | Ramp (<500 m) | 0.30 | 0.16 | 1.89 | 0.064 | 1.73 |
| Count | Restaurants (< 350 m) | 0.65 | 0.20 | 3.17 | 0.003 | 2.99 |
| **NO<sub>x</sub> (adj. R<sup>2</sup> = 0.71, p<0.001)** |  |  |  |  |  |  |
|  | Constant | 0.07 | 0.17 | 0.39 | 0.697 |  |
| Total length | Primary collector (<250 m) | 0.84 | 0.27 | 3.13 | 0.003 | 2.37 |
|  | Local (<500 m) | 0.80 | 0.24 | 3.31 | 0.002 | 1.92 |
|  | Major arterial (<500 m) | 0.69 | 0.26 | 2.67 | 0.010 | 2.18 |
|  | Ramp (<500 m) | 1.07 | 0.26 | 4.04 | 0.000 | 2.27 |
| Count | Restaurants (< 100 m) | 0.89 | 0.27 | 3.28 | 0.002 | 2.42 |
|  | Restaurants (< 350 m) | 0.64 | 0.22 | 2.89 | 0.006 | 1.59 |
| Proximity | Major arterial | 1.12 | 0.33 | 3.40 | 0.001 | 3.52 |
| **O<sub>3</sub> (adj. R<sup>2</sup> = 0.53, p<0.001)** |  |  |  |  |  |  |
|  | Constant | 0.08 | 0.24 | 0.33 | 0.743 |  |
| Total length | Highway (<100 m) | -1.76 | 0.29 | -6.15 | 0.000 | 1.42 |
|  | Minor arterial (<350 m) | -2.44 | 0.51 | -4.76 | 0.000 | 3.28 |
|  | Local (<500 m) | -1.37 | 0.30 | -4.62 | 0.000 | 1.52 |
| Count | Restaurants (<250 m) | -0.69 | 0.31 | -2.25 | 0.029 | 1.63 |
| Area | NDVI (500 m) | 0.46 | 0.26 | 1.78 | 0.082 | 1.19 |
| Proximity | Highway | -1.11 | 0.33 | -3.35 | 0.002 | 1.90 |

All models displayed strong associations with cumulative length of roads by type, with these predictors explaining nearly 30% of the variation in the pollutant concentrations. Interestingly, total length of all roads was eliminated at all buffer distances, but that for specific types—most commonly, local and major arterial—remained significant. The number of restaurants in 100 m, 250 m, or 250 m were positively associated for NO, NO_2_, and NO_x_, and negatively associated for O_3_. O_3_ had inverse associations with cumulative occurrence of and proximity to roadways as well as number of restaurants, but strongly increased with NDVI within 500 m. The only significant negative correlation in the remaining models was between NO and NDVI within 50 m. but strongly increased with NDVI within 500 m. No other greenness parameters (e.g. leaf biomass and leaf area) were retained in the models.

## Discussion

This study demonstrated that 50-70% of the spatial heterogeneity in NO, NO_2_, NO_x_, and O_3_—all known to vary at the neighborhood and urban levels—can be explained by land-use parameters characterizing proximity to and cumulative occurrence of local emission sources as traffic and restaurants. The results add to the body of evidence that hyperlocal differences in pollutant concentrations, even within acceptable regulatory levels, remain persistent. Models developed to fit 2019 annual averages predicted the spatial heterogeneity in pollutant levels measured in 2020, suggesting that the associations are robust to annual differences in meteorology and pollutant levels. This performance is especially significant since traffic and emission patterns may have differed in the latter portion of 2020 due to city-wide lockdowns. The models developed in this work illustrate that differences within small spatial extents that appear relatively homogeneous— the study domain in Louisville was predominantly composed on single-family residences—can moderate patterns in air pollution.

We observed that while the presence of expressways strongly influenced proximal pollutant mixing ratios of NO and O_3_, the significant road types for NO_2_ and NO_x_ were smaller and designed to conduct traffic to the residential neighborhoods. The predictors identified by models in the present study have been detected in previous LUR modeling of intra-urban NO_2_ and O_3_ (8,9,21,23,34). A key difference, however, is that land cover variables such as industrial or commercial use land were also significant in these investigations. That such parameters were not retained in the models developed for this study’s domain is consistent with the relatively low diversity in its land cover. It should also be noted that the modeling extents in the aforementioned studies spanned major cities in North America and Europe. Consistent with the observations in Louisville, investigators from the ESCAPE project, which developed LUR models for NO_2_ and NO_x_ for 36 areas in Europe, observed that predictors differed with both spatial scales and complexities of the built environment (27).

We did not include meteorological parameters such as wind direction of a sampling location relative to major emission sources as done by Jerrett *et al*. because I identified no major sources within the study domain with zones of representation incorporating a significant fraction (>10%) of the sampling sites (30). The effects due to the interstate highway were captured by proximity parameters, but it is possible that persistent effects due to emissions from the adjacent airport or industrial point sources were neglected. This could also explain why the models developed using annual average concentrations had lower predictive capability for season averages: the models could not capture effects due to seasonal shifts in prevailing wind patterns. Adding these parameters could increase the variance in annual average pollutant levels explained by the models and improve the fitting for datasets collected over small temporal scales.

A limitation of modeling at this spatial scale is that data to directly quantify traffic count such as annual average daily traffic, traffic intensity, or traffic volume for highways and major roads, are typically not available at the spatial resolution required. The observation from this study that ambient pollutant concentrations are associated with hyperlocal characteristics in roadways warrants that parameters quantifying other land use and land cover features, particularly greenness, also be augmented. In this work, I quantified greenness by area (fraction of area within a given buffer occupied by parks, mean NDVI/leaf biomass/leaf area within a buffer); it is also likely that other predictors of land use directly or inversely captured greenness (e.g. larger area for industrial use or roadways). It is possible, however, that more nuanced treatment of vegetative indices (stratification of leaf area or leaf biomass by percentiles, stratification by trees through simplified group of species, and interaction terms of these two strata) could capture changes in local air quality due to either structure (large canopies impede air flow and elevate pollutant levels) or interaction (pollutant uptake differ across plant species) (41–44). Recent investigations have found urban air pollution abatement can vary by plant species (45). In addition to addressing the seasonal instability of the models and limited predictive power for the model in fitting O_3_, such enhancements would allow for assessments of health effects in the current study domain— collaborators have already determined that residential greenness is associated with exposure to volatile organic compounds and cardiovascular disease risk in a larger area (Jefferson County) which encompasses the extent of my analysis (46,47).

In summary, this work has two main implications: first, hyperlocal assessments in environmental epidemiology must consider fine-scale geographic features as these may influence chronic exposures to urban air pollution; second, the heterogeneity in exposures, while beyond the spatial resolutions of regulatory monitoring sites, can be captured by small-area, fixed-site networks using passive samplers. Empirical models to robustly predict exposures across various time scales, however, will require measured data at the corresponding time scales.

## Data Availability

All data produced in the present study are available upon reasonable request to the authors

## Acknowledgements

This work was supported by the National Institute of Health [NIH 1R01ES29846-01, PI A. Bhatnagar, PhD, FAHA, University of Louisville] and the Nature Conservancy. Dr. Jason Su (University of California, Berkeley) identified sampling locations using the location-allocation method. Mr. Rick Strehl (University of Louisville) deployed and retrieved the passive samplers.

## Supplementary material

## Notes

### Competing Interest Statement

The authors have declared no competing interest.

